# Impact of antenatal iron deficiency on maternal heart function-A hypothesis-generating translational study

**DOI:** 10.64898/2026.03.06.26347784

**Authors:** M Vera-Aviles, C Sinaida, S.N Kabir, M.D Christodoulou, S Kasner, A Frost, L Heather, C Aye, A Arulalagan, F Samuels, B Raman, P Leeson, M Nair, S Lakhal-Littleton

## Abstract

**Background and aims:** Iron deficiency (ID) and myocardial iron depletion (MID) are causally linked to heart failure (HF) in the general population and in preclinical models. ID is common amongst pregnant women, but its impact on cardiac adaptations to pregnancy is unknown. This study examines that impact, and its potential relevance to peripartum cardiomyopathy (PPCM).

**Methods:** We provided female mice with iron-replete or iron-deficient diets, and monitored cardiac function and morphology longitudinally in pregnancy and postpartum. In women with no HF (n=64), we explored the associations between antenatal iron parameters and echocardiographic parameters in late pregnancy and at 6-12 months postpartum. We also performed a case (n=55), control (n=170) study comparing iron markers and assessing their association with PPCM risk.

**Results:** In mice, ID prevented postpartum reversal of pregnancy-induced hypertrophy, reduced postpartum LVEF, and caused profound MID. In women with no HF, low hepcidin, high transferrin and low serum iron were respectively associated with higher LVESV, lower LVEF and higher CMR T1-mapping (lower myocardial iron) in postpartum. In the PPCM study, serum iron, hepcidin and haemoglobin were significantly lower in cases than controls, and were independently associated with risk of PPCM. Mechanistically, myocardial proteomics revealed that ID caused sustained postpartum activation of pyruvate dehydrogenase kinase 4, a master cardiometabolic switch enzyme with a well-recognised role in HF.

**Conclusions:** This study links antenatal maternal ID to postpartum systolic dysfunction, and implicates MID and cardiometabolic switching as potential mechanisms. It suggests these links may potentially contribute to the pathophysiology of PPCM.

**TRANSLATIONAL PERSPECTIVE:**

- Iron deficiency (ID), even without anaemia, is linked to risk of incident HF in the general population and to worse outcomes in those with pre-existing HF. Preclinical studies demonstrated a causal role for myocardial iron depletion (MID) in this context.
- ID is common during pregnancy, but its impact on the maternal heart, and its adaptations to the haemodynamic stress of pregnancy are unknown
- By revealing that antenatal ID is associated with postpartum systolic dysfunction and with PPCM, this study points to ID as a novel risk factor for maternal HF.
- By revealing the role of MID in this context, this study highlights the potential of intravenous iron therapies, which we have previously shown to raise myocardial iron, to reduce risk of maternal HF.

GRAPHICAL ABSTRACT

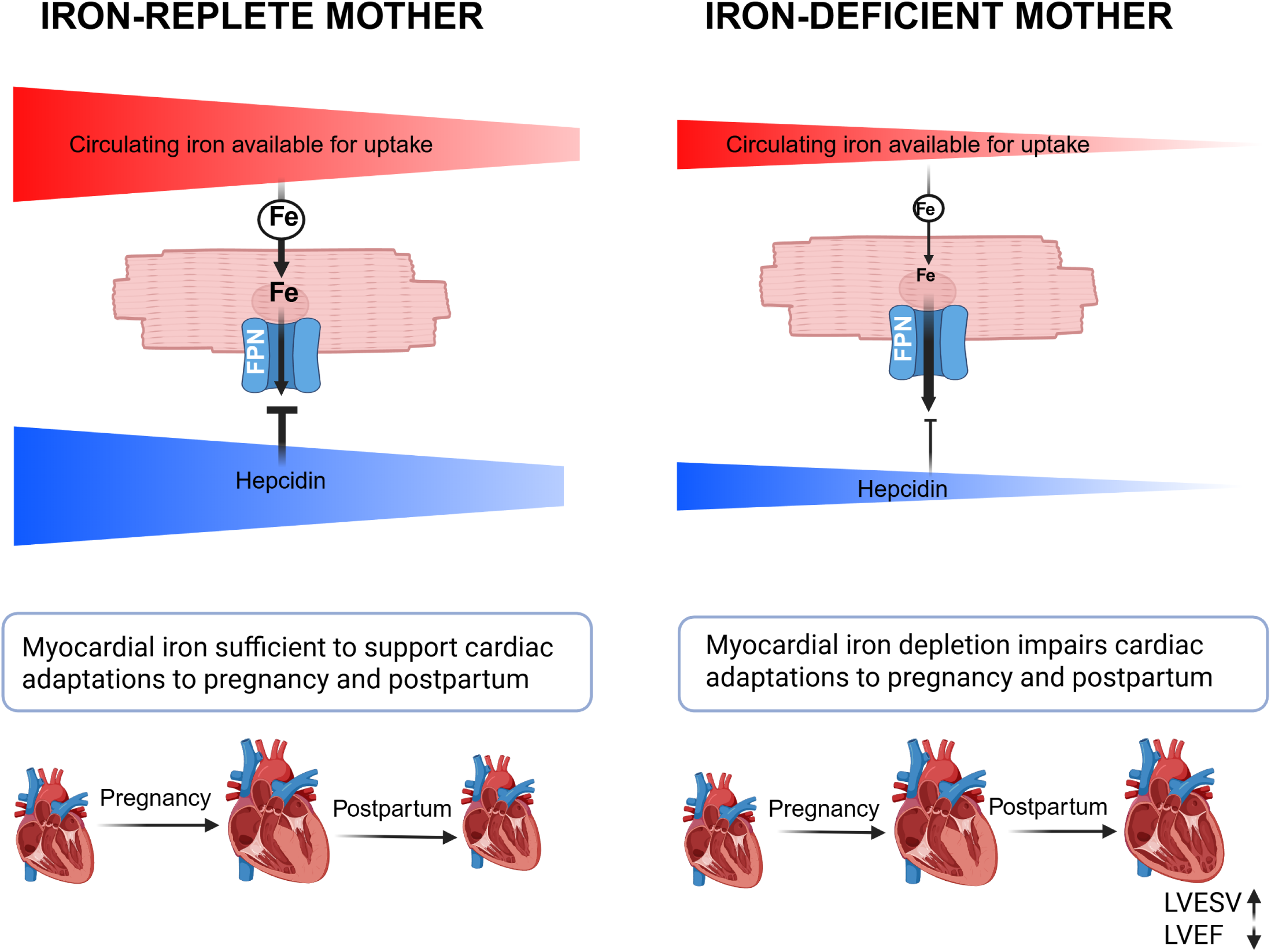

## INTRODUCTION

Despite clinical guidelines for screening and supplementing iron levels in the antenatal period, iron deficiency (ID) remains highly prevalent amongst pregnant women^1^. Indeed, an estimated 37% women receiving standard oral iron supplementation, and 77% of non-iron supplemented women develop ID by the third trimster^2-4^. This reflects heightened demand for iron, needed to support both the expansion of maternal blood volume and rapid fetal growth^5^. This demand results in gradual depletion of maternal iron levels, manifesting as reduced circulating iron availability (denoted by a reduction in serum iron and transferrin saturation Tsat) and depletion of maternal iron stores (denoted by reduction in serum ferritin and increase in serum transferrin)^6-8^. Pregnancy is also accompanied by suppression of maternal hepcidin, the central hormone of iron homeostasis. The aim of hepcidin suppression is to increase plasma iron availably by relieving the block on iron absorption and mobilisation from stores, though the impact of hepcidin suppression is limited in women with depleted iron stores ^6-8^.

Outside the context of pregnancy, a diagnosis of ID, based on low ferritin and or low Tsat is associated with incident risk of heart failure (HF), and with worse outcomes in patients with acute or chronic HF, independently of anaemia^9-11^. Low hepcidin is also associated with higher mortality in patients with HF^12-14^. Previous discoveries by this team provided a mechanistic framework for how the combination of ID and of low hepcidin impair heart function. We discovered that, beyond controlling iron absorption and mobilisation, hepcidin is also directly required for controlling iron export from cardiomyocytes. We showed that uncontrolled iron export from cardiomyocytes leads to myocardial iron depletion (MID), impaired cardiac energetics, and eventually HF with reduced ejection fraction (HFrEF)^15^.

Pregnancy poses a physiological challenge for the maternal heart. The maternal heart adapts through left ventricular hypertrophic remodelling that peaks in the third trimester, and is normally reversed by 1 month postpartum^16-19^. Maladaptation to the physiological stress of pregnancy is a hallmark of peripartum cardiomyopathy PPCM, a rare form of de-novo HF manifesting in late pregnancy or early postpartum period^20^.

Despite the well-recognised links between ID and HF in the general population, the impact of maternal ID on maternal heart adaptations to pregnancy, and on postpartum reverse-remodelling remain unexplored.

This study addresses this question by combining observations from pregnant women with a clinically-recapitulative mouse model of pregnancy and ID. In pregnant mice, we induce ID to determine its impact on cardiac function in pregnancy and postpartum, quantify iron levels directly in the myocardial tissue, and perform proteomic analysis to further uncover the mechanisms linking ID to altered heart function. In women without a diagnosis of HF, we examine the relationship between maternal iron markers, and heart morphology and contractility during pregnancy and postpartum. We also examine the relationship between maternal iron markers and myocardial iron levels, estimated by CMR T1 mapping. Finally, we explore the relevance of these findings to the pathophysiology of PPCM through a case-control study.

## METHODS

### Mouse model of pregnancy and iron deficiency

All animal procedures were compliant with the UK Home Office Mice Scientific Procedures Act 1986 (licence# P84F13B1B) and approved by the University of Oxford Medical Sciences Division Ethical Review Committee. B6;FVB female mice were used. To safeguard against severe anaemia, female mice were maintained on an iron-replete diet containing 200 parts per million (ppm) iron, from birth until 5 weeks of age, after which they were randomised to continue on the iron-replete diet or switch to an iron-restricted diet containing 5 ppm iron (Teklad TD.99397). Randomisation was carried out using the Rand() function in excel. At 11 weeks of age, half the females were mated with B6;FVB males, and inspected daily for a plug. They were then maintained on their respective diet for the remainder of the experiment. Non-pregnant females maintained on the respective diets were also used as controls. Each data point represents a single mouse. No mice were excluded from the analysis.

### MRI assessment of cardiac function in mice

Cardiac function and morphology were evaluated by Cine MRI at baseline (2±1 gestational age), third trimester (17+/1 days gestational age), then at 1 week, 3 weeks and 6 weeks postpartum. For the Cine MRI scan, mice were anaesthetized with 2% isofluorane in O_2_ and positioned supine in a purpose-built cradle. ECG electrodes were inserted into the forepaws, a respiration loop was taped across the chest and heart signals were monitored using a custom-built physiological motion gating device. The cradle was lowered into a vertical-bore, 11.7 T MR system with a 40 mm birdcage coil (Rapid Biomedical, Würzburg, Germany) and visualised using a Bruker console running Paravision 2.1.1. A stack of contiguous 1 mm thick true short-axis ECG and respiration-gated cine-FLASH images were acquired. The entire in vivo imaging protocol was performed in approximately 60 min. Image analysis was performed using ImageJ (NIH Image, Bethesda, MD). Left ventricular volumes and ejection fractions were calculated from the stack of cine images. For pregnant females, weight at baseline was used as denominator in calculating heart/body weight ratio.

### Assessment of systemic and cardiac iron homeostasis in mice

Serum iron indices were measured in blood collected via the tail vein using the ABX-Pentra C400 system (Horiba) as detailed in our previous work^15^. Total cardiac iron levels were evaluated in mouse hearts by induced couple plasma mass spectrometry (ICP-MS) as per our previous work^15^. Liver and cardiac hepcidin gene expression was evaluated by Taqman gene expression assays (Applied Biosystems). Cardiac ferroportin protein was detected in formalin-fixed paraffin-embedded sections, using rabbit polyclonal anti-mouse FPN antibody (NBP1-21502, Novus Biologicals) at 1/200 dilution, followed by Alexa 647- conjugated anti-rabbit secondary antibody (ab150075, Abcam), diluted at 1:1000. Following nuclear counter-stain with 4′,6-diamidino-2-phenylindole (DAPI), coverslips were visualised using a Fluoview FV1000 confocal microscope (Olympus). All immunostaining experiments included an isotype control, followed by Alexa 647- conjugated anti-rabbit secondary antibody.

### Proteomic analysis in mouse myocardial tissue

Details are provided in supplemental methods.

### Exploratory study in women with no diagnosis of HF (CAREFOL-HT cohort)

Clinical samples and data were obtained from 64 participants in the ongoing Clinical Randomised Antenatal Study to Characterise Key Roles of Tetrahydrofolate in Hypertensive Pregnancies (CAREFOL-HT), a multi-centre, double-blind, randomised controlled trial. All participants included in this analysis were recruited from the Women’s Centre at Oxford University Hospitals NHS Foundation Trust (UK) between September 2021 and August 2024. Participants were aged 18–45 years. Of the 64 women included in our analysis, 25 were from the preeclampsia arm of the study. These had a clinician-confirmed diagnosis of preeclampsia. Preeclampsia was defined as gestational hypertension (systolic blood pressure ≥140 mmHg and/or diastolic blood pressure ≥90 mmHg, presenting after 20 weeks’ gestation on repeated measurements), together with either (a) proteinuria (urine protein-to-creatinine ratio ≥30 mg/mmol) and/or (b) evidence of uteroplacental dysfunction. A normotensive comparator cohort was recruited comprising women with blood pressure <140/90 mmHg throughout pregnancy and fewer than two moderate risk factors for hypertensive disease in pregnancy, according to the National Institute for Health and Care Excellence (NICE) guideline for the management of hypertension in pregnancy^21^. The trial was prospectively registered at ClinicalTrials.gov (NCT05434195) and overseen by an independent trial steering committee and data safety monitoring committee. Ethical and research governance approval was obtained from the Wales Research Ethics Committee (21/WA/0169; IRAS Project ID: 295830). Baseline demographic data, including age, sex, ethnicity, and antenatal data from 12 weeks’ gestation, were collected.

### Iron indices in CAREFOL-HT cohort

We evaluated iron indices in blood samples collected at the first study visit, corresponding to median gestational age [IQR] of 28+2 [27+4, 29+3] weeks+days. Serum ferritin, transferrin and iron concentrations were determined using the ABX-Pentra C400 system (Horiba) as detailed in our previous work^15^. Serum hepcidin was determined by Ella Simple Plex ELISA system (Biotechne Catalog # SPCKB-PS-000984).

### Echocardiographic parameters in CAREFOL-HT cohort

We evaluated the relationship between iron markers collected at the first study visit, and echocardiographic parameters collected in late pregnancy, coinciding with median gestational age [IQR] of 36 [35+3, 36+4], and in postpartum, corresponding with 6-12 months after delivery.

Standardised transthoracic echocardiography was performed using Philips EPIQ 7C or CX50 ultrasound systems equipped with two-dimensional phased-array transducers (Philips Healthcare). Image acquisition was performed with participants in the left lateral decubitus position, and data collection followed the British Society of Echocardiography (BSE) minimum dataset^22^.

Echocardiographic analyses were performed offline using IntelliSpace Cardiovascular (ISCV), version 4.1 (Philips Healthcare), by observers blinded to clinical data. All measurements were taken in accordance with the BSE minimum dataset. Body surface area (BSA) was calculated using the Mosteller equation.

Assessment of left ventricular (LV) structure included two-dimensional measurements of LV wall thickness and calculation of LV mass, which was indexed to BSA. Relative wall thickness was calculated as the sum of interventricular septal thickness and posterior wall thickness at end-diastole divided by the left ventricular internal diameter at end-diastole, providing an index of concentric LV remodelling. LV volumes and systolic function were assessed using the biplane Simpson’s method to derive LV end-diastolic and end-systolic volumes, and left ventricular ejection fraction.

Assessment of LV diastolic function included pulsed-wave Doppler evaluation of mitral inflow to derive early (E) and late (A) diastolic peak velocities and calculation of the E/A ratio. Tissue Doppler imaging was used to measure early diastolic myocardial velocities (e′) at the septal and lateral mitral annulus, with E/e′ ratios calculated as an index of LV filling pressure. Left atrial volumes were quantified using standard biplane methods and indexed to BSA.

### CMR in CAREFOL-HT cohort

Magnetic resonance imaging was performed at the postpartum visit using a 3.0 Tesla Prisma Scanner (Siemens Medical Solutions, Erlangen, Germany) at the Oxford Center for Clinical Magnetic Resonance Research (John Radcliffe Hospital, Oxford, United Kingdom).

Quantitative myocardial tissue characterisation was performed using cvi42 by experienced analysts blinded to clinical data. Native T1 mapping analysis was performed by drawing myocardial contours on mid ventricular short axis T1 maps, with care taken to exclude blood pool and artefact. Average myocardial native T1 values were generated for each participant. Map quality was assessed using R2 maps derived from the shortened modified Look Locker inversion recovery fitting to ensure robust data fidelity. Myocardial T2 mapping analysis was conducted on mid ventricular short axis images using the Siemens MyoMaps T2 prepared balanced steady state free precession sequence. Myocardial contours were manually defined and mean myocardial T2 values were calculated, excluding regions of artefact and blood pool signal, to assess myocardial oedema or inflammation.

### PPCM case control study

This case-control study was nested within an ongoing case-control study of acute HF in pregnant and postpartum women in India undertaken through the Maternal and perinatal Health Research collaboration, India (MaatHRI)^23^. Ten selected public and private hospitals in three states in India enrolled women presenting with HF at any point during pregnancy, childbirth, and up to 12 months after childbirth. Data were collected between February 2019 and January 2023. The criteria for inclusion in the study were: 1) being at least 18 years old, 2) being pregnant or within 1 year postpartum, and 3) being willing and able to give informed consent for participating in the study. The control group comprised pregnant women not diagnosed with any cardiac problems, who had given birth within two days of the case presenting at the hospital. In this study a random subset of the control group was compared to a subset who were diagnosed with PPCM based on clinical history and echocardiography findings according to the diagnostic criteria suggested by the European Society of Cardiology (ESC):

i. symptoms of HF developing towards the end of pregnancy or in the months following delivery;
ii. no other identifiable cause of HF;
iii. no recognizable structural heart disease prior to the last month of pregnancy; and
iv. a reduced LVEF (<45%).

Blood samples were collected non-fasted from all women at the time of enrolment in the study. Serum concentrations of ferritin were measured using a Chemiluminescence Immunoassay (Siemens ADVIA Centaur) following the manufacturer’s instructions. Serum iron levels were measured by Spectrophotometry, while Tsat values were calculated from measurements of serum iron and total iron binding capacity (measured by Spectrophotometry; Siemens Atellica). Levels of hepcidin were measured using an enzyme-linked immunosorbent assay (EIA).

## STATISTICAL ANALYSIS

Normality and equal variance were tested using the Shapiro–Wilk test and Levene test, respectively, and choice of parametric vs non-parametric tests applied accordingly. Significance was set at p<0.05.

Preclinical data are reported as mean±standard error of the mean (s.e.m). Multiple group comparisons were drawn by two-way ANOVA, or mixed effects analysis to account for missing data, followed by Sidak’s post hoc test.

Clinical data are reported as median [Interquartile range].

In the CAREFOL-HT cohort, correlations were drawn using Pearson’s or Spearman’s test. The results of these correlations are reported in heatmaps. Significant correlations are denoted by an * to show p<0.05. Correction for multiple comparisons was not performed due to the exploratory nature of this study.

In the PPCM case-control study, iron markers were compared between women with PPCM and control women using Wilcoxon rank sum tests. The association between each iron marker and PPCM was then assessed in separate univariable and multivariable logistic regression models. A directed acyclic graph (DAG) was developed to identify potential confounders of the association between ID and PPCM. Maternal education level, hypertensive disorders in pregnancy, maternal age, body mass index (BMI), socioeconomic status, postpartum blood loss, parity and duration of gestation were considered as risk factors for the condition, but maternal education was the only one identified as a potential confounder of the association. The main effect of the iron markers on risk of PPCM wase estimated for either every 1 unit change from median (for Tsat, and Haemoglobin), or 10 units change from median (for hepcidin and serum iron).

## RESULTS

### ID in mice impairs reverse-remodelling of pregnancy-induced cardiac hypertrophy and reduces postpartum LV contractile function

To test the impact of ID on pregnancy-related cardiac remodelling and its postpartum reversal, we used a mouse model of pregnancy. Females were given iron-deficient (ID) or iron-replete (IR) diets for 6 weeks before pregnancy and maintained on these diets throughout. We used MRI to perform a longitudinal assessment of cardiac morphology and contractility at baseline, 3^rd^ trimester then at 1, 3 and 6 weeks postpartum (PP).

In iron-replete females, heart/body weight ratio increased in the third trimester relative to baseline, but returned to baseline by 6 weeks postpartum (Fig1A). These changes recapitulate the well-recognised processes of hypertrophic growth, and its postpartum reversal in the human heart. The magnitude of hypertrophic remodelling during pregnancy was not significantly different between iron-replete and iron-deficient females. However, hypertrophic remodelling was not reversed in iron-deficient females in the postpartum period (1A). Similarly, LVEF was not different between iron-replete and iron-deficient females during pregnancy, but LVEF was significantly lower in iron-deficient females, both at 1 week and at 6 weeks postpartum (1B).

**Figure 1.**
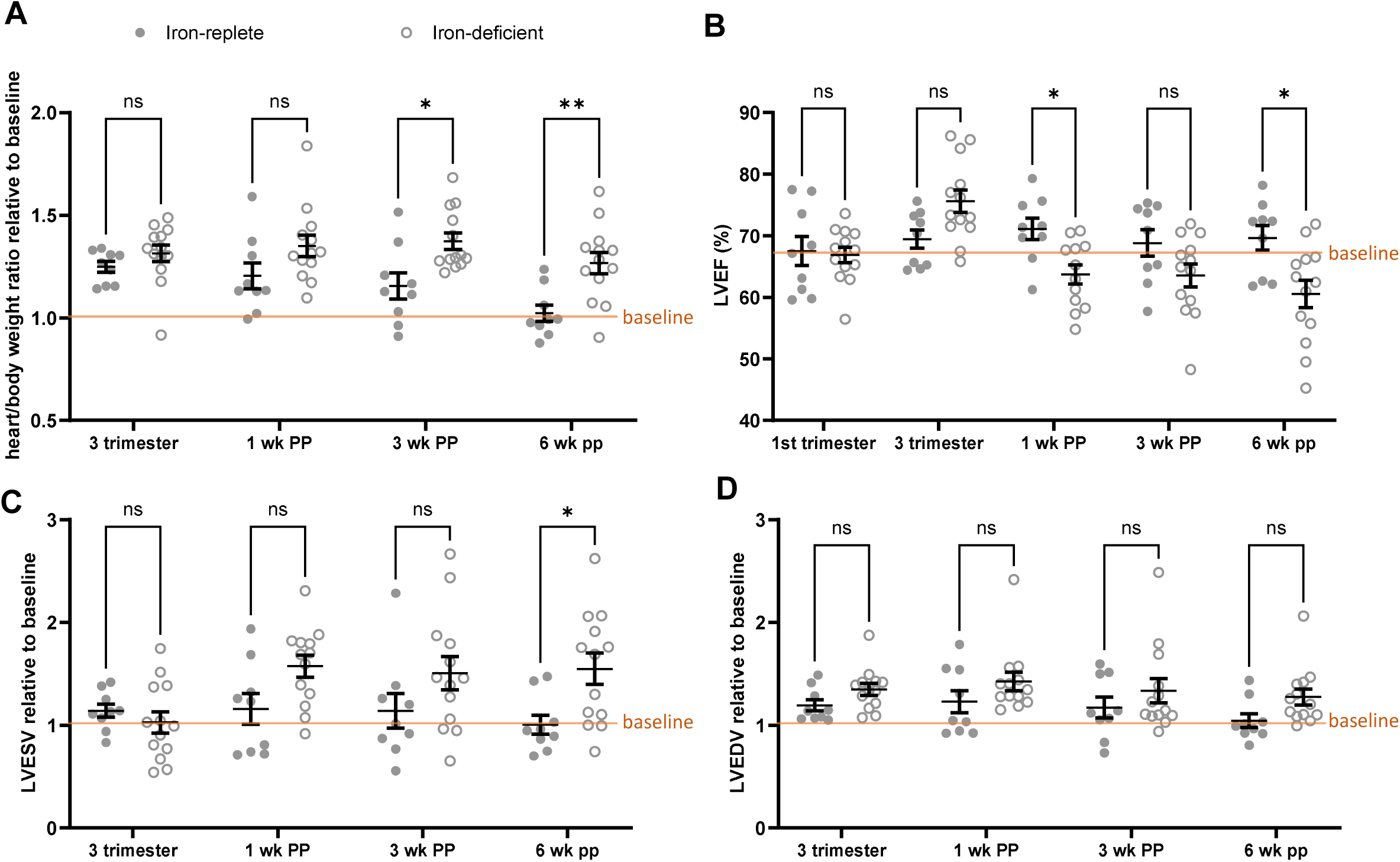
Iron deficiency in mice reduces postpartum contractile function and delays reverse-remodelling of the maternal heart. Longitudinal MRI assessment of changes in heart/body weight ratio, LVEF, LVESV and LVEDV in iron-replete (n=9) and iron-deficient (n=13) mice. Data were collected at baseline (1^st^ trimester), 3^rd^ trimester, 1 week postpartum (1WK PP), 3 weeks postpartum (3wk PP) and 6 weeks postpartum (6wk PP). A. Changes in heart to body weight ratio relative to baseline B. LVEF at all timepoints C. Changes in LVESV relative to baseline D. Changes in LVEDV relative to baseline Two-way repeated measure ANOVA with Šidák correction for multiple comparisons. *p<0.05, **p<0.01, ns=not significant

LV remodelling was not different between iron-deficient and iron-replete females, during pregnancy and early post-partum, but LVESV was significantly higher in iron-deficient females than iron-replete females at 6 weeks post-partum (1C). LVEDV was not different between iron-replete and iron-deficient females at any timepoint (1D).

In the absence of pregnancy, we did not observe any differences in cardiac parameters between iron-replete and iron-deficient females at the respective timepoints (supplemental Fig 1).

We confirmed that females maintained on an iron-deficient diet had reduced serum iron levels (2A), Tsat (2B) and ferritin (2C), compared to those maintained on iron-replete diet, regardless of pregnancy and timepoint. Progression of pregnancy from first to third trimester was accompanied by a drop in serum iron, Tsat and ferritin levels. In the postpartum period, these markers recovered relative to third trimester in iron-replete females, but failed to recover in iron-deficient females.

Haemoglobin levels trended towards being lower in iron-deficient vs iron-replete females, though this was not statistically significant (Fig2D). In iron-deficient females, progression of pregnancy from the first to third trimester was accompanied by a significant decline in haemoglobin, which did not recover in the postpartum period.

**Figure 2.**
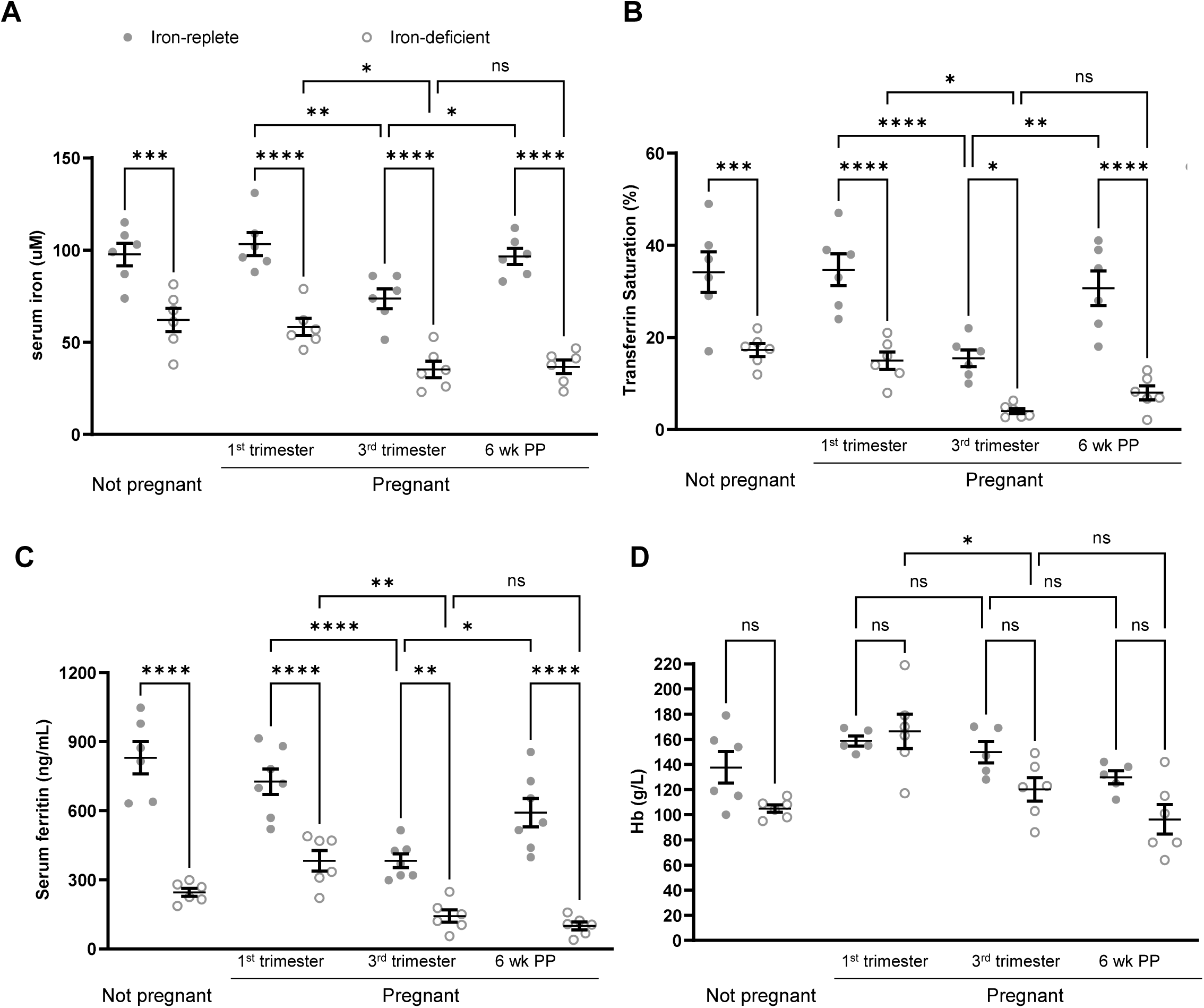
Impact of iron-deficient diet and pregnancy on iron markers in mice. Terminal collection of blood was performed IN iron-replete and iron-deficient female mice at 1^st^ trimester, 3^rd^ trimester and 6 weeks postpartum (6wk PP). Non-pregnant female mice were used as controls. A. Serum iron concentration B. Transferrin Saturation (Tsat) C. Serum ferritin D. Haemoglobin Two-way ANOVA with Šidák correction for multiple comparisons. *p<0.05, **p<0.01, ***p<0.001, ****p<0.0001, ns= not significant

These findings demonstrate that maternal ID impairs postpartum reverse-remodelling of LV hypertrophy, resulting in impaired LV systolic function.

### The combination of ID and pregnancy leads to profound hepcidin suppression, elevation of myocardial ferroportin, and myocardial iron depletion

We previously discovered that cardiomyocytes express the iron exporter ferroportin (FPN) and that this FPN is under control of hepcidin, both of cardiac and of hepatic origins^15,24^. We also showed that loss of this control leads to unregulated iron export, myocardial iron depletion and heart failure with reduced ejection fraction HFrEF in mice^15,24.^ Increased myocardial FPN expression together with lowered myocardial iron content have since also been observed in biopsies from failing human hearts^25,26^. To explore this paradigm in the context of pregnancy, we evaluated the expression of hepatic (Fig3A) and cardiac hepcidin (Fig3B), cardiac FPN (Fig3C) and quantified myocardial iron levels (Fig3D). Progression of pregnancy in iron-replete females was accompanied by suppression of liver hepcidin expression, but this expression recovered in postpartum. This is consistent with hepcidin changes in human pregnancy. In the absence of pregnancy, ID also reduced liver hepcidin expression, consistent with the normal homeostatic regulation of hepatic hepcidin by transferrin saturation. However, the combination of ID and pregnancy resulted in the most profound suppression of liver hepcidin in the third trimester, which failed to recover in the postpartum period (Fig 3A).

**Figure 3.**
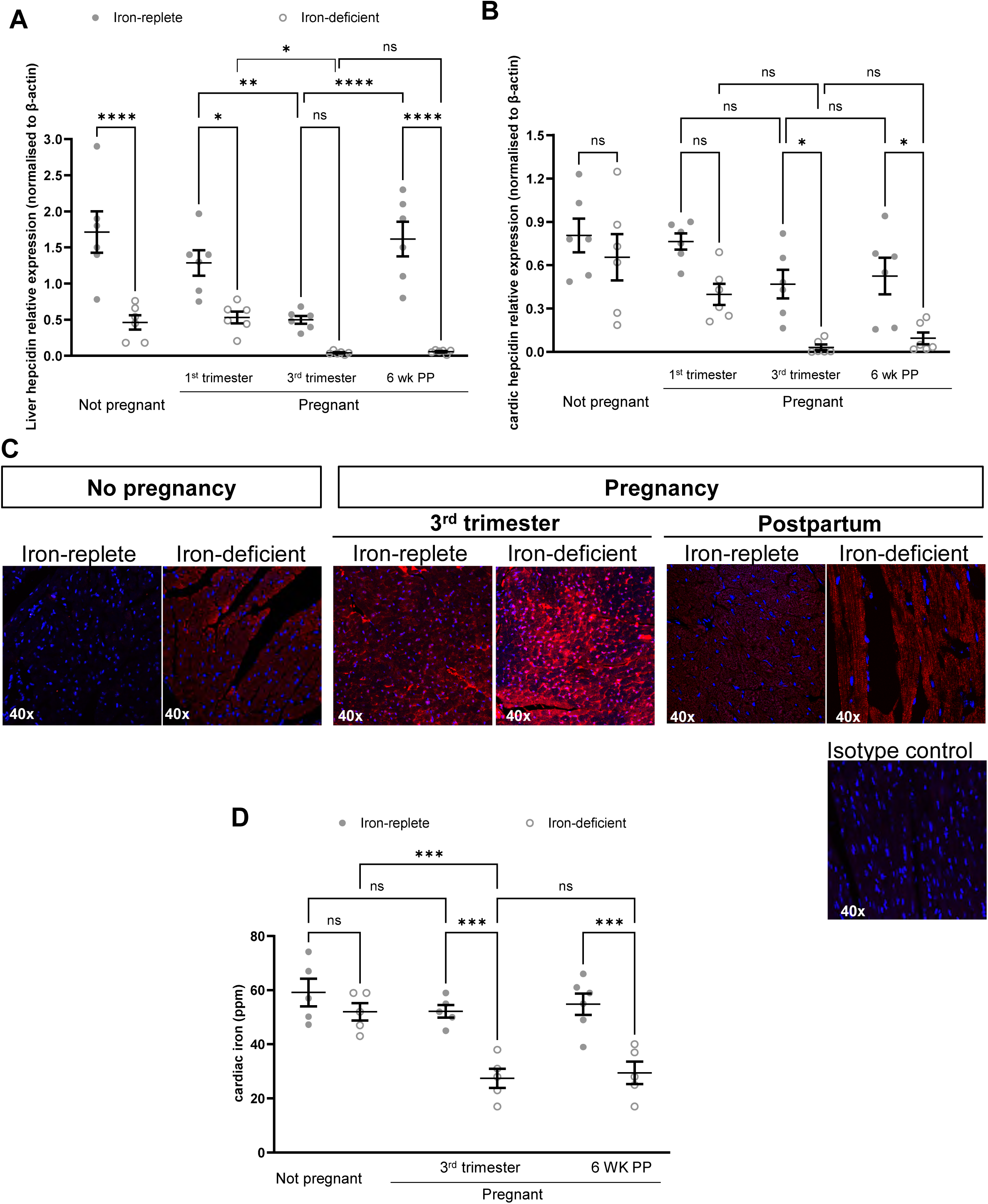
The combination of iron deficiency and pregnancy leads to profound hepcidin suppression, elevation of myocardial ferroportin, and myocardial iron depletion. Terminal collection of hearts and livers was performed at in iron-replete and iron-deficient female mice at 1^st^ trimester, 3^rd^ trimester and 6 weeks postpartum (6wk PP). Non-pregnant female mice were used as controls. A. Relative hepcidin gene expression in the liver B. Relative hepcidin gene expression in the heart C. Representative images of ferroportin immunostaining (red) in the heart. Nuclei are blue. D. Quantitation of cardiac iron content, ppm= parts per million Two-way ANOVA with Šidák correction for multiple comparisons. *p<0.05, **p<0.01, ***p<0.001, ****p<0.0001, ns= not significant

Cardiac hepcidin was not significantly affected by either ID or pregnancy alone, but profoundly suppressed by the combination of pregnancy and ID, in the third trimester, failing to recover in the postpartum period (Fig3B)

In agreement with the profound suppression of cardiac and hepatic hepcidins, the combination of pregnancy and ID also resulted in the most profound increase in cardiac FPN levels in the 3^rd^ trimester (3C). This was accompanied by depleted myocardial iron content in pregnant iron-deficient females, which did not recover in the postpartum period (3D).

These data demonstrate that ID and pregnancy act synergistically to suppress maternal hepcidin and drive myocardial iron depletion (MID).

### Antenatal maternal ID during is associated with altered cardiac morphology and function in pregnancy and postpartum

Next, we sought to explore the translational value of preclinical findings using data from women. The study included 64 women with no prior diagnosis of HF, with a median age at consent of 33 [29.8-36.0] years. Of these, 25 (39.1%) had preeclampsia during the index pregnancy, and 3 (3.1%) had hypertension before pregnancy. Median gestational age [IQR] at visit 1 was 28+2 [27+4, 29+3] weeks+days. Baseline demographics are shown in table 1.

**Table 1.** Demographics and baseline characteristics of women in CAREFOL-HT study.

|  |  |
| --- | --- |
| **Age at consent, median [IQR] (years)** | 33.0 [29.8, 36.0] |
| **Ethnicity, n (%)** |  |
| Caucasian | 53 (82.8%) |
| Black | 5 (7.8%) |
| South Asian | 3 (4.7%) |
| East Asian | 3 (4.7%) |
| **First trimester BMI mean [IQR]** | 24.5 [22.3, 28.6] |
| **Parity n (%)** |  |
| Nulliparous | 40 (62.5%) |
| Multiparous | 18 (28.1%) |
| **Gravida n (%)** |  |
| 1 | 29 (45.3%) |
| 2 | 20 (31.3%) |
| 3 | 9 (14.1%) |
| 4 | 4 (6.3%) |
| 5 | 1 (1.6%) |
| 6 | 1 (1.6%) |
| **Hypertension during pregnancy n (%)** |  |
| No | 39 (60.9%) |
| Yes | 25 (39.1%) |
| **Hypertension before pregnancy n (%)** |  |
| No | 62 (96.9%) |
| Yes | 2 (3.1%) |
| **Gestational diabetes n (%)** |  |
| No | 62 (96.9%) |
| Yes | 2 (3.1%) |
| **Diabetes before pregnancy n (%)** |  |
| No | 62 (96.9%) |
| Yes | 2 (3.1%) |
| **Mode of delivery, n (%)** |  |
| Spontaneous vaginal delivery | 24 (37.5%) |
| Instrumental | 7 (10.9%) |
| Emergency C section | 22 (34.4%) |
| Elective C section | 11 (17.2%) |
| **Smoking status n (%)** |  |
| Never smoked | 57 (89.1%) |
| Quit>3 months ago | 4 (6.3%) |
| Quit <3 months ago | 2 (3.1%) |
| Current smoker | 1 (1.6%) |
| **Gestational age at visit 1, median [IQR] (weeks+days)** | 28+2 [27+4, 29+3] |
| **Gestational age at visit 2, median [IQR] (weeks+days)** | 36 [35+7, 36+4] |
| Missing | 11 (17.2%) |
| **Gestation age at delivery, median [IQR] (weeks+days)** | 39 [34+6, 39+5] |
| **Time of postpartum visit from delivery, median [IQR] (months)** | 8.53 [7.65, 10.8] |
| **Iron supplementation during pregnancy, n(%)** |  |
| Oral (Ferrous sulphate or ferrous fumarate) | 7 (10.9%) |
| Intravenous (Ferinject) | 2 (3.1%) |
| None | 55 (86%) |
| **Serum Iron at visit 1 (uM), median [IQR]** | 17.2 [11.4, 22.6] |
| Missing, n (%) | 14 (21.9%) |
| **Tsat at visit 1 (%), median [IQR]** | 25.0 [15.1, 32.8] |
| Missing, n (%) | 14 (21.9%) |
| **Hepcidin at visit 1 (pg/mL), median [IQR]** | 597.5 [127.5, 2384.8] |
| Missing, n (%) | 14 (21.9%) |
| **Ferritin at visit 1 (ng/mL), median [IQR]** | 22.5 [9.5, 34.2] |
| Missing, n (%) | 15 (23.4%) |
| **Transferrin at visit 1 (g/L), median [IQR]** | 3.1 [2.4-3.3] |
| Missing, n (%) | 14 (21.9%) |
| **Haemoglobin at visit 1 (g/L), median [IQR]** | 114.0 [108.0, 121.0] |
| Missing, n (%) | 0 (0%) |
| **HCT at visit 1 (%), median [IQR]** | 34.6 [32.9, 36.6] |
| Missing, n (%) | 0 (0%) |

We performed an exploratory analysis of the correlations between iron markers at visit 1 and parameters of cardiac morphology and contractility measured in late pregnancy (Fig4A) and postpartum (Fig4B). We also evaluated the relationship between iron markers and CMR native T1 mapping, and native T2, as surrogate markers of myocardial iron and OF oedema/inflammation respectively. The r values for all correlations are summarised in heatmaps (Fig4).

**Figure 4.**
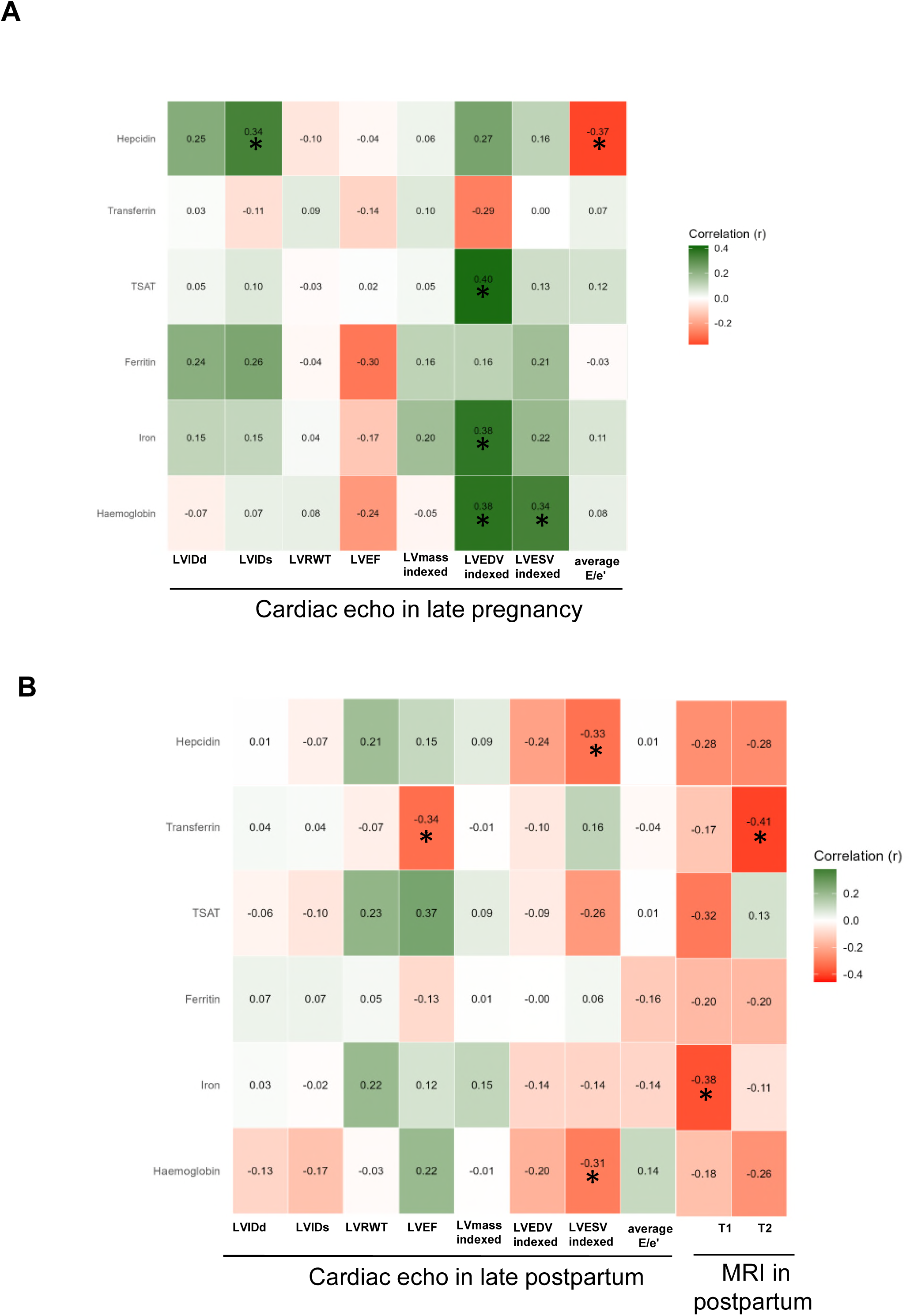
Correlation between maternal iron markers and cardiac function and morphology in CAREFOL-HT cohort. A. Correlation between maternal iron markers at visit 1 and cardiac parameters at late pregnancy B. Correlation between maternal iron markers at visit 1 and cardiac parameters, and CMR T1 and T2 native mapping at postpartum LVIDd= Left Ventricular Internal Diameter at end-diastole LVIDs= Left Ventricular Internal Diameter at end-systole LVRWT= Left ventricular relative wall thickness LVEF= Left ventricular ejection fraction LV mass indexed= Left ventricular mass indexed LVEDV indexed= Left ventricular end-diastolic volume indexed LVESV indexed= Left ventricular end-systolic volume indexed Average E/e’= ratio of mitral peak early filling velocity to early diastolic mitral annular velocity Spearman’s rank correlation test, r values are shown. *p<0.05, **p<0.01

Low Hepcidin levels at visit 1 correlated significantly with lower Left ventricular internal dimension in systole (LVIDs) and higher ratio of early mitral inflow velocity (E) to early diastolic mitral annular velocity (e’) (Average E-e) at visit 2 and with larger Left ventricular end-systolic volume (LVESV) in postpartum.

Higher transferrin at visit 1 correlated significantly with lower LVEF and lower myocardial T2 at postpartum.

Lower Tsat at visit 1 correlated significantly with lower LVEDV at visit 2 but not in postpartum.

Lower serum iron at visit 1 correlated significantly with lower LVEDV at visit 2 and with higher myocardial T1 in postpartum.

Finally, lower Haemoglobin at visit 1 correlated significantly with lower LVEDV and LVESV in at visit 2 but higher LVESV in postpartum.

These exploratory analyses link antenatal maternal ID to smaller LV dimensions and increased LV filling pressure in late pregnancy, consistent with co-centric hypertrophy. On the other hand, antenatal maternal ID during pregnancy appears to be associated with reduced LV systolic function (lower LVEF, higher LVESV) in postpartum, consistent with systolic dysfunction. The correlation between low serum iron and high myocardial T1 suggests that maternal ID may also be associated with myocardial iron depletion (MID).

Due to the low numbers of paired variables in women with pre-eclampsia, sub-group analysis was not possible for any of the variables at visit 2, and only possible for some of the variables at postpartum. These showed that the correlation between transferrin and LVEF was mainly driven by the normotensive group (r=-0.44, p=0.028 in normotensive vs r=-0.10, p=0.71 in pre-eclampsia). The correlation between transferrin and CMR T2 was significant or trended towards significance in both groups (r=-0.41, p=0.038 in normotensive, vs r=-0.49, p=0.085 in pre-eclampsia). The correlation between serum iron and CMR T1, and between hepcidin and LVESV were no longer significant in the subgroup analysis.

These data mirror preclinical findings and further support the notion that maternal ID has a negative impact on postpartum heart function.

### Maternal ID is associated with PPCM

Having observed a link between maternal ID and sub-clinical systolic dysfunction, we sought to explore if this link may be relevant to the pathophysiology of PPCM, a form of dilated cardiomyopathy, characterised by severe systolic dysfunction presenting typically between the last month of pregnancy and up to six months postpartum. We conducted a nested case-control study, involving 55 women diagnosed with PPCM and 170 control postpartum women with no cardiac problems. Baseline characteristics of controls and cases are shown in table 2.

**Table 2:**
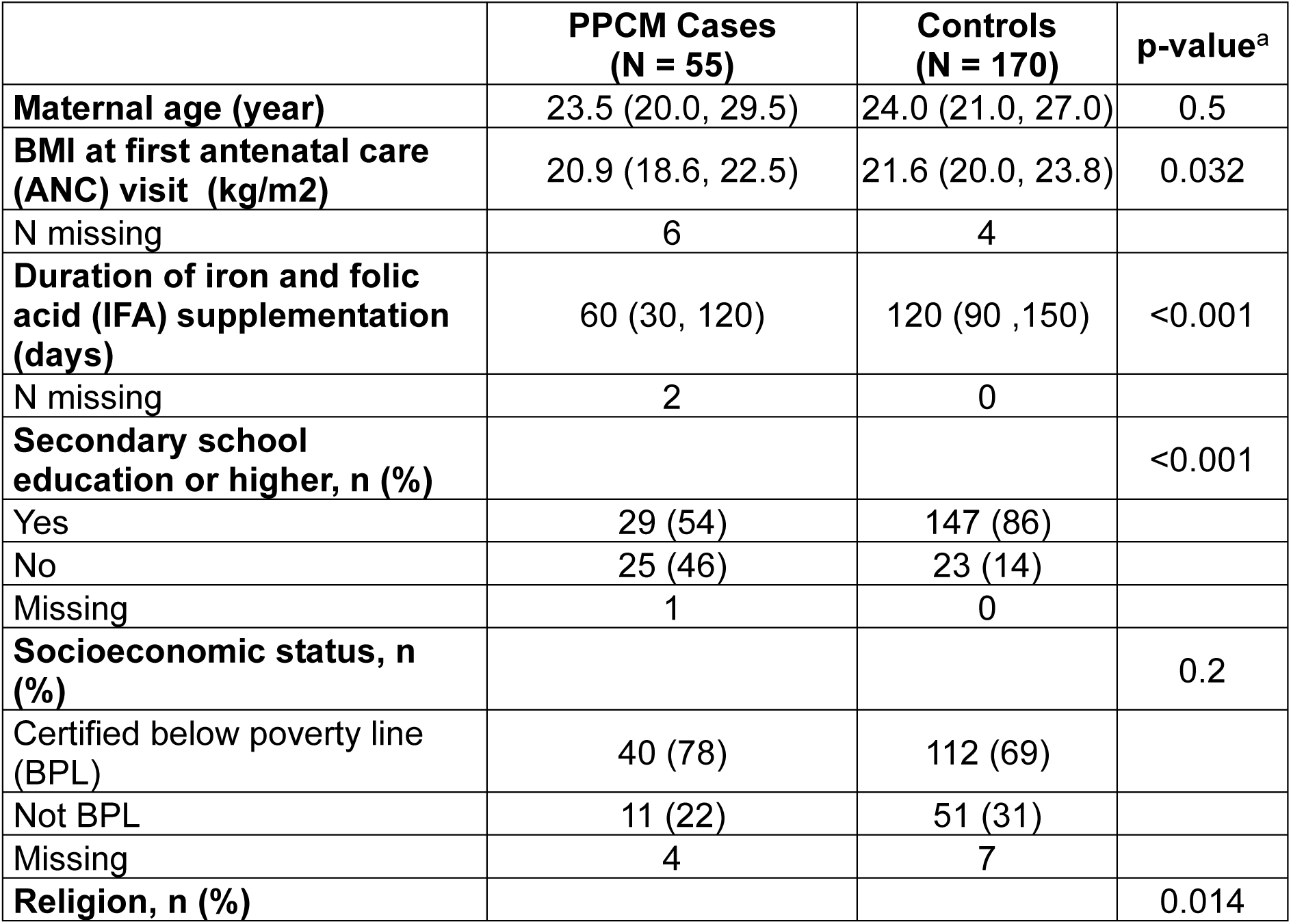
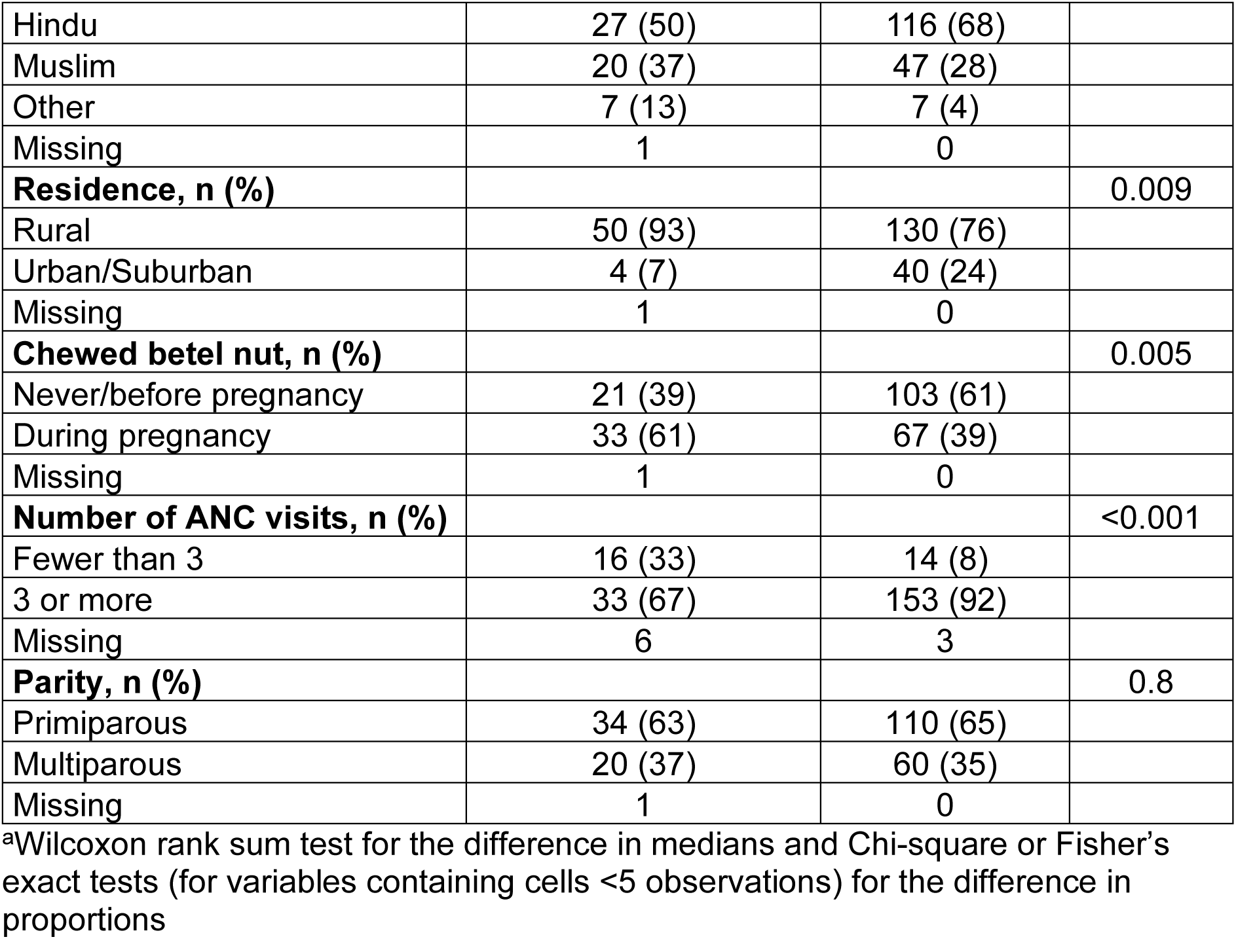
Baseline characteristics on cases and controls.

|  | **PPCM Cases**<br>**(N = 55)** | **Controls**<br>**(N = 170)** | **p-value<sup>a</sup>** |
| --- | --- | --- | --- |
| **Maternal age (year)** | 23.5 (20.0, 29.5) | 24.0 (21.0, 27.0) | 0.5 |
| **BMI at first antenatal care**<br>**(ANC) visit (kg/m<sup>2</sup>)** | 20.9 (18.6, 22.5) | 21.6 (20.0, 23.8) | 0.032 |
| N missing | 6 | 4 |  |
| **Duration of iron and folic**<br>**acid (IFA) supplementation**<br>**(days)** | 60 (30, 120) | 120 (90, 150) | <0.001 |
| N missing | 2 | 0 |  |
| **Secondary school**<br>**education or higher, n (%)** |  |  | <0.001 |
| Yes | 29 (54) | 147 (86) |  |
| No | 25 (46) | 23 (14) |  |
| Missing | 1 | 0 |  |
| **Socioeconomic status, n**<br>**(%)** |  |  | 0.2 |
| Certified below poverty line<br>(BPL) | 40 (78) | 112 (69) |  |
| Not BPL | 11 (22) | 51 (31) |  |
| Missing | 4 | 7 |  |
| **Religion, n (%)** |  |  | 0.014 |
| Hindu | 27 (50) | 116 (68) |  |
| Muslim | 20 (37) | 47 (28) |  |
| Other | 7 (13) | 7 (4) |  |
| Missing | 1 | 0 |  |
| **Residence, n (%)** |  |  | 0.009 |
| Rural | 50 (93) | 130 (76) |  |
| Urban/Suburban | 4 (7) | 40 (24) |  |
| Missing | 1 | 0 |  |
| **Chewed betel nut, n (%)** |  |  | 0.005 |
| Never/before pregnancy | 21 (39) | 103 (61) |  |
| During pregnancy | 33 (61) | 67 (39) |  |
| Missing | 1 | 0 |  |
| **Number of ANC visits, n (%)** |  |  | <0.001 |
| Fewer than 3 | 16 (33) | 14 (8) |  |
| 3 or more | 33 (67) | 153 (92) |  |
| Missing | 6 | 3 |  |
| **Parity, n (%)** |  |  | 0.8 |
| Primiparous | 34 (63) | 110 (65) |  |
| Multiparous | 20 (37) | 60 (35) |  |
| Missing | 1 | 0 |  |

<sup>a</sup>Wilcoxon rank sum test for the difference in medians and Chi-square or Fisher's exact tests (for variables containing cells <5 observations) for the difference in proportions

First, we compared iron markers between the two groups. Women with PPCM had significantly lower concentrations of serum iron, hepcidin and lower haemoglobin than control women. TSAT levels were not significantly different between the women with PPCM and controls (Table 3).

**Table 3:** Iron biomarkers in PPCM cases and controls.

|  | **PPCM Cases**<br>**N = 55** | **Controls**<br>**N = 170** | **p-value<sup>a</sup>** |
| --- | --- | --- | --- |
| Serum iron (ug/dL), median (IQR) | 43 (23, 82) | 65 (44, 105) | <0.002 |
| N missing | 11 | 1 |  |
| TSAT (%), median (IQR) | 12 (7,25) | 16 (10,25) | 0.11 |
| N missing | 11 | 4 |  |
| Hepcidin (ng/mL), median (IQR) | 2 (1, 12) | 13 (3, 30) | <0.001 |
| N missing | 15 | 16 |  |
| Hb (g/dL), median (IQR) | 9.2 (7.90, 11.00) | 10.50 (9.60, 11.40) | 0.002 |
| N missing | 2 | 0 |  |

<sup>a</sup>Wilcoxon rank-sum test

We then performed multivariate regression analysis adjusting for identified confounders (education) and found that the odds of PPCM increased by 40% with every 10 unit decrease from the median concentration of hepcidin (aOR 1.40, 95% CI: 1.10, 1.70), by 11% with every 10 units decrease from the median concentration of serum iron (aOR 1.11, 95% CI: 1.04, 1.20). The odds of PPCM were not significantly associated with Tsat or haemoglobin. (Fig 5).

**Figure 5.**
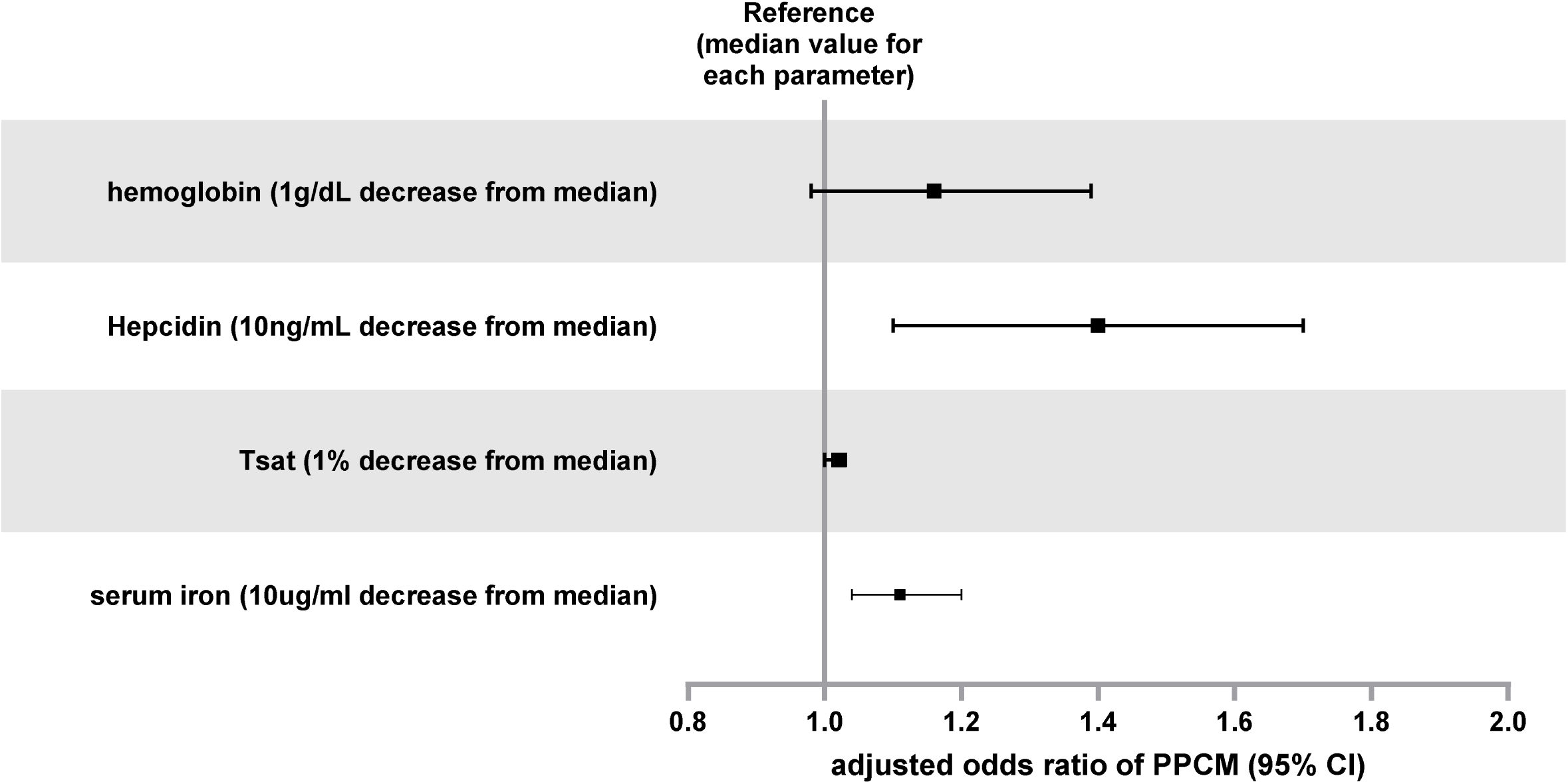
Maternal iron depletion is associated with peripartum cardiomyopathy. Multivariable logistic regression model was used to estimate adjusted odds ratio of PPCM for every 1g/L decrease from median haemoglobin, 10ng/mL decrease from median hepcidin, 1% decrease from median Tsat, and 10ug/ml decrease from median iron.

### ID in mice hinders postpartum reverse-remodelling of myocardial energetics

Finally, we sought to further explore the mechanisms linking ID with postpartum systolic dysfunction. We performed a proteomic analysis of myocardial tissues from mice. We identified 306 proteins that are significantly changed in postpartum relative to 3^rd^ trimester, of which 106 were significantly upregulated and 200 were significantly downregulated during the postpartum period (Fig6A). We also identified 93 proteins that were significantly different between iron-deficient and iron-replete hearts in the postpartum period, including, 58 that were downregulated by ID, and 35 that were upregulated by ID (Fig6B). Importantly, expression of the uptake protein transferrin receptor (TfRC) was increased while expression of iron storage proteins ferritin heavy (FTH1) and light chains (FTL1) were decreased in hearts from iron-deficient vs hearts from iron-replete mice, consistent with the expected homeostatic responses to MID, driven by Iron Regulatory Proteins (IRPs). There were 11 proteins that were significantly altered both in postpartum and by ID (Fig6C). Of these, only 3 proteins were changed in opposing directions. Cathepsin J and pyruvate dehydrogenase kinase (PDK4) were downregulated during postpartum in iron-replete hearts, but were higher in iron-deficient hearts than iron-replete hearts, while Semaphorin 4B (SEMA4B) was upregulated in postpartum hearts but was lower in hearts from iron-deficient hearts than iron-replete hearts.

**Figure 6.**
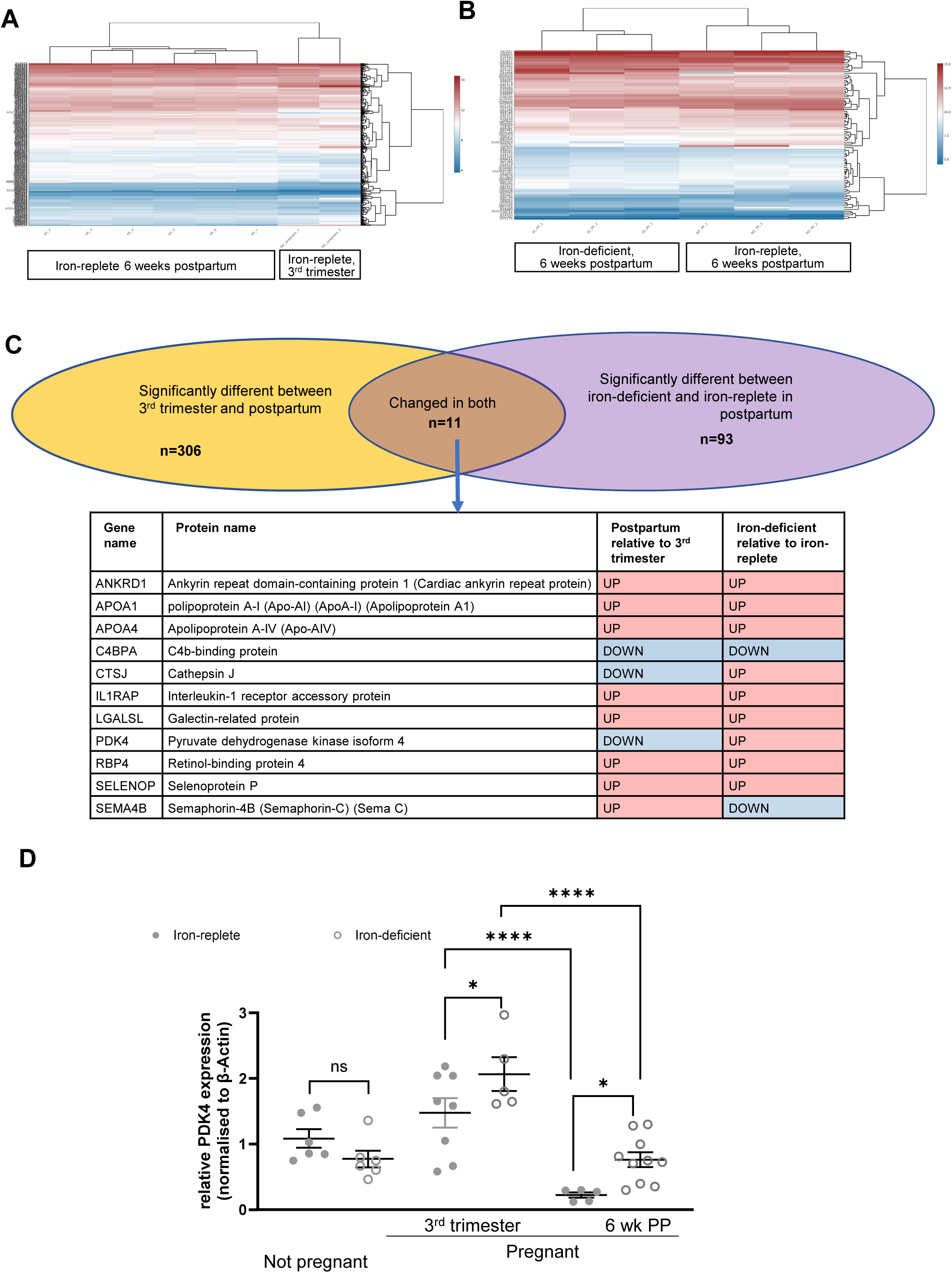
Iron deficiency hinders postpartum reverse-remodelling of myocardial metabolism. A. Heatmap representing myocardial proteins that are significantly altered between the 3^rd^ trimester of pregnancy and 6 weeks postpartum B. Heatmap representing myocardial proteins that are significantly different between iron-replete and iron-deficient females at 6 weeks postpartum C. Venn diagram summarizing the number of proteins that are both significantly changed during postpartum and affected by iron deficiency. D. Relative gene expression of pyruvate dehydrogenase kinase (PDK4) in hearts of iron-replete and iron-deficient females at 3^rd^ trimester and 6 weeks postpartum. Non-pregnant females controls are also shown. Two-way ANOVA with Šidák correction for multiple comparisons. *p<0.05, ****p<0.0001, ns= not significant

Outside the context of pregnancy, elevated myocardial PDK4 expression has been causally implicated in cardiac hypertrophy and heart failure^27-30^. PDK4 is a mitochondrial enzyme that inhibits the pyruvate dehydrogenase complex, thus reducing the conversion of pyruvate that is produced from of glucose, into acetyl-CoA. In this manner, higher expression of PDK4 limits the ability of the heart to utilise glucose, and increases its reliance on fatty acids oxidation as an energetic fuel^27,28^. Consistent with this, we noted that higher PDK4 levels in postpartum hearts from iron-deficient females were accompanied by lower levels of ATP-dependent 6-phosphofructokinase (PFK-A), the enzyme that catalyses the first committed step of glycolysis (Fig6B).

To further understand the links between pregnancy, ID and myocardial PDK4, we evaluated its gene expression in greater detail. We found that ID alone did not significantly affect myocardial pdk4 gene expression. There was a significant decline in pdk4 gene expression in the postpartum period compared to the third trimester, consistent with the proteomic data. Both in the third trimester and in postpartum, pdk4 gene expression was significantly higher in iron-deficient hearts relative to iron-replete hearts (Fig6D).

These data suggest that postpartum reverse-remodelling of the maternal heart is accompanied by remodelling of myocardial energetics towards greater use of glucose, but that ID promotes continued reliance on fatty acid oxidation.

## DISCUSSION

Outside the context of pregnancy, ID, even without anaemia is associated with risk of incident HF in the general population and with worse outcomes in those with pre-existing heart disease^9-14^. ID is especially prevalent during pregnancy and the perinatal period, coinciding with significant haemodynamic stress for the maternal heart^2-8^. Despite this, its impact on the maternal heart has remained unexplored, until now. The present study is the first report to link maternal ID to systolic dysfunction in the postpartum period. It implicates myocardial iron depletion and cardiometabolic switching as the underlying mechanisms. It also suggests that these links may contribute to the pathophysiology of PPCM.

Our exploratory study in pregnant women with no known diagnosis of HF found an association between markers of depleted iron status and reduced LV systolic function in postpartum. The preclinical model of ID recapitulated this association, thus serving to demonstrate the causal role of ID. The preclinical model also revealed the synergetic effects of ID and pregnancy in suppressing maternal hepcidin, increasing cardiac ferroportin (FPN) and depleting myocardial iron. This myocardial iron depletion mirrored the association between low serum iron levels and higher myocardial T1 in women. Previous work by this team was the first to identify myocardial iron depletion as a mechanistic link between ID and HF^15^. Indeed, we showed that mice harbouring targeted deletion of myocardial hepcidin, or targeted substitution of myocardial FPN with a hepcidin-resistant form, had markedly upregulated myocardial FPN, increased myocardial export, myocardial iron depletion, and a progressive decline in systolic function towards HFrEF^15^. Together, these discoveries point to myocardial iron depletion as a likely contributor to systolic dysfunction in the postpartum heart. This work underscores the need for further research using larger human cohorts to fully elucidate the impact of maternal ID on maternal heart function in pregnancy and postpartum, as well as its prognostic relationship with the risk of HF in the long-term.

Further mechanistic exploration using proteomic analysis revealed that expression of PDK4, a master metabolic enzyme that promotes fatty acids over glucose as energetic fuel, is markedly higher in the late pregnancy than in postpartum. This observation is consistent with a well-recognised role fatty acids, which are more energy-dense than glucose, in supporting the heightened energetic demands of hypertrophic remodelling and increased cardiac output in late pregnancy^27,28^. Notably, we found that PDK4 was downregulated in postpartum, but that ID was associated with sustainedly high PDK4 expression in the postpartum period. Sustainedly elevated PDK4 expression is a significant finding in this context, because elevated myocardial PDK4 expression has been causally linked to progression from hypertrophic cardiomyopathy to HF in humans and in preclinical models^29,30^. This work suggests that sustained PDK4 expression in iron-deficient hearts may well contribute to systolic dysfunction in the postpartum period. Further research is needed to understand the interplay between myocardial iron content, PDK4 expression and maternal adaptations to pregnancy and postpartum.

Finally, our finding that maternal ID was associated with sub-clinical changes in cardiac function in women with no HF prompted us to explore its potential relevance to the pathophysiology of PPCM. The aetiology of PPCM remains poorly understood, with evidence thus far pointing to mechanisms involving oxidative stress and abnormal cleavage of prolactin^20^. Our observation that markers of maternal ID are also associated with risk of PPCM adds an additional and novel mechanism, that could plausibly interact with other risk factors and disease pathways to engender this acute and severe form of systolic dysfunction. Of relevance, several studies have shown that myocardial native T1 is elevated in women with PPCM compared to women without cardiac complications and furthermore that higher native T1 identified PPCM cases that did not recover LVEF over time^31,32^. While native myocardial T1 is commonly regarded as an indicator of oedema and fibrosis, it is in fact directly correlated with myocardial iron levels. Indeed, UK Biobank studies found myocardial T1 to be strongly determined by gene variants in iron homeostatic pathways including those in the hepcidin pathway^33, 34^. Thus, myocardial iron depletion may also be mechanistically involved in the pathophysiology of PPCM. Further research is needed to formally evaluate the role of myocardial iron in PPCM, e.g. using T2* mapping.

The hypotheses generated by this translational study could have important clinical implications. They point to antenatal ID as a potential risk factor for PPCM in the short-term, and for HF in the long-term. They also underscore the limitations of current guidelines on the management of ID in pregnancy^1^. Indeed, these guidelines are anaemia-centric, and do not currently advocate for active monitoring of other iron markers, despite the fact that anaemia is often the last consequence of ID. Our work suggests that measuring hepcidin, transferrin and serum iron could identify women with greatest unmet need for iron in the heart, and consequently greater risk of cardiac complications. Furthermore, guidelines on iron supplementation advocate using oral tablets to treat anaemia, with intravenous iron therapies only recommended from the second trimester in women who do not tolerate or respond to oral iron. Whether these supplementation approaches are sufficient to prevent myocardial iron depletion in pregnancy remains unknown. However, our own previous work has shown that the intravenous iron drug ferric carboxymaltose (FCM) raises myocardial iron content^35^. One implication is that IV iron therapy could be used in pregnancy to rapidly replete myocardial iron and ensure the heart can sustain the haemodynamic stress of pregnancy and postpartum reverse-remodelling.

### Study strengths and limitations

The strength of this a translational study is that it integrates clinical observation with preclinical models to aid both causal inference and mechanistic understanding. Several markers of maternal iron homeostasis are measured including hepcidin in order to fully capture maternal iron status. Evaluation of myocardial T1 in the clinical study, together with direct quantitation of myocardial iron in the preclinical model strengthen the case for myocardial iron depletion being relevant to systolic dysfunction. In the preclinical study, the effect of ID on the maternal heart is systematically and thoroughly investigated through longitudinal MRI in pregnancy and postpartum. Non-pregnant iron-deficient controls are included to further tease out the effects of ID from those of pregnancy. The study has a number of limitations. The small size of the exploratory pregnancy cohort precludes comprehensive sub-group analysis and any multivariate regression analysis to fully account for potential confounders, such as inflammation that can influence both iron homeostasis and reflect cardiovascular dysfunction. The results of that study are purely correlative, serving to provide novel hypotheses for further testing. In the PPCM study, controls were not matched to PCCM cases by gestational age, as these were recruited within 48h postpartum for logistical feasibility.

## DECLARATIONS

S.L.-L. reports receipt of previous research funding from Vifor Pharma, personal honoraria on a lecture from Pharmacosmos and consultancy fees from Disc Medicine and ScholarRock.

## DATA AVAILABILITY

The data underlying this article will be shared on reasonable request to the corresponding author.

## HUMAN SUBJECTS

All human research in this article complies with the Declaration of Helsinki, and has been approved by the locally appointed ethics committee. All patients gave informed consent.

