## supplemental figure 1 for "Impact of antenatal iron deficiency on maternal heart function-A hypothesis-generating translational study"

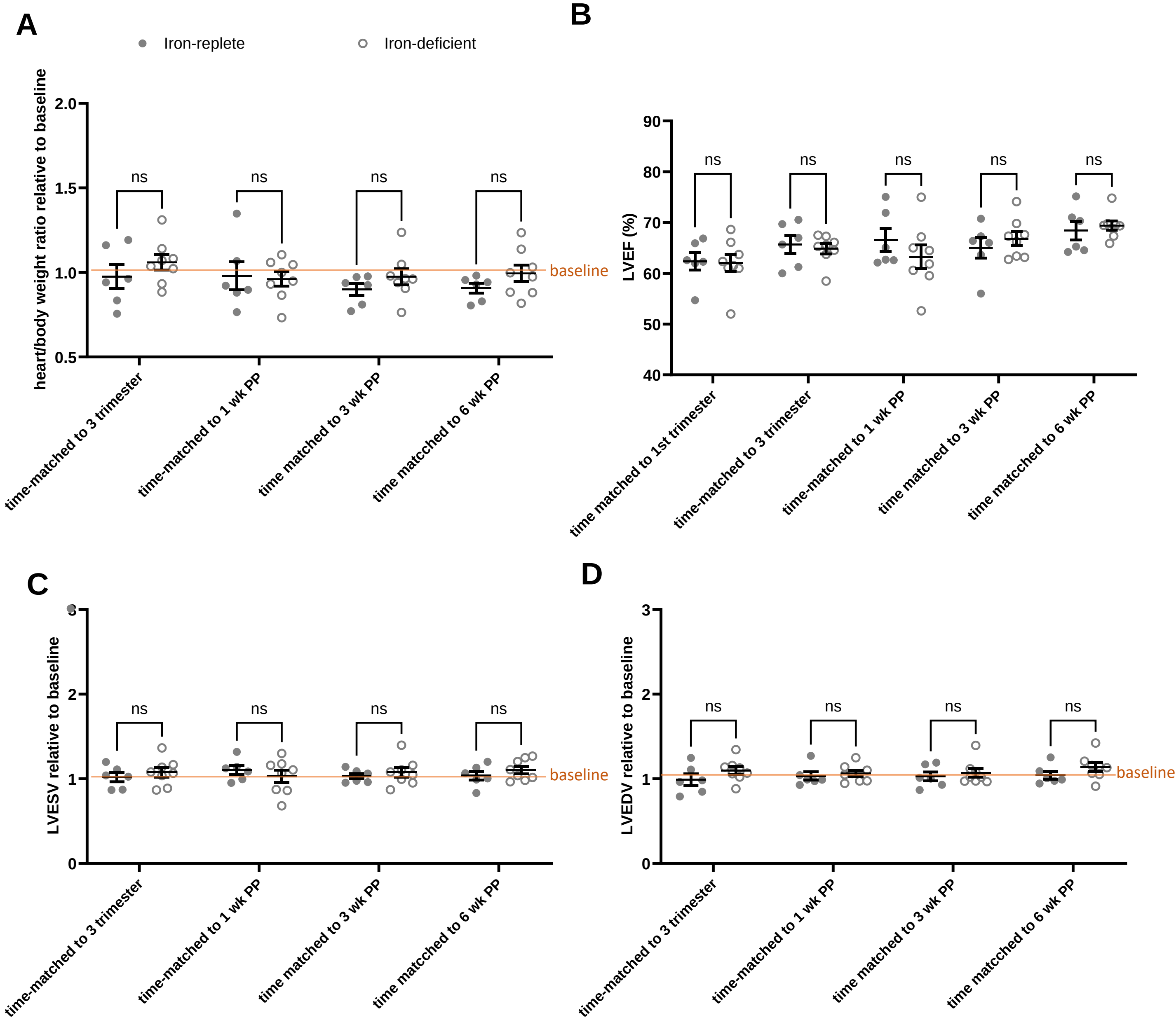

**Supplemental Figure 1-Iron deficiency alone does not affect heart function in female mice.** Longitudinal MRI assessment of changes in heart/body weight ratio, LVEF, LVESV and LVEDV in iron-replete (n=6) and iron-deficient (n=8) mice. Data were collected at timepoints to match those in pregnant females, 1st trimester, 3rd trimester, 1 week postpartum (1WK PP), 3 weeks postpartum (3wk PP) and 6 weeks postpartum (6wk PP).

A. Changes in heart to body weight ratio relative to baseline
B. LVEF at all timepoints
C. Changes in LVESV relative to baseline
D. Changes in LVEDV relative to baseline

Two-way repeated measure ANOVA with Šidák correction for multiple comparisons. ns=not significant
