## supplemental methods for "Impact of antenatal iron deficiency on maternal heart function-A hypothesis-generating translational study"

**Proteomic analysis of mouse myocardial tissue**

**Sample preparation** 75 µL of Lysis buffer (50 mM Tris-HCl, pH 8.5, 5 mM EDTA, 150 mM NaCl, 10 mM KCl, 1% Triton X-100, 5 mM sodium fluoride (NaF), 5 mM beta-glycerophosphate, 1 mM Na-orthovanadate, containing Roche complete protease inhibitor) was added to each sample along with a 3 mm tungsten carbide bead (Qiagen). Samples were treated twice in the TissueLyser II homogenizer (Qiagen) going from 3-30 Hz in 60 seconds. The beads were removed and samples were transferred to clean Eppendorf LoBind tubes and incubated for 2h at -4°C. The samples were centrifuged at 15.000 g for 20 min at 4°C and supernatant precipitated with 25% acetone and left at -20°C O.N. The samples were centrifuged at 400g for 1.5 min and the pellets were left to dry. Samples were added 20 µL Lysis buffer (6M Guanidinium Hydrochloride, 10 mM TCEP, 40 mM CAA, 50 mM HEPES pH 8.5) and treated twice in the TissueLyser II homogenizer (Qiagen) going from 3-30 Hz in 60 seconds. The beads were removed and samples were transferred to clean Eppendorf LoBind tubes and boiled at 95°C for 5 minutes, followed by sonication on high for 5x 60 seconds on and 30 seconds off in a Bioruptor Pico sonication water bath (Diagenode) at 4°C. Samples were centrifuged at 18,000 RCF for 10 minutes, and supernatants were transferred to clean Eppendorf Protein LoBind tubes. Protein concentrations were determined by BCA Rapid Gold (Thermo Scientific) and 20 µg protein was taken forward for digestion. Sample volume was normalized between samples with additional lysis buffer and diluted 3x with digestion buffer (10% Acetonitrile in 50 mM HEPES pH 8.5) and 400 ng of LysC (MS-grade Wako) was added. Samples were incubated for 3 hours at 37°C, shaking at 750 RPM. After LysC digestion, samples were diluted to a final 10x in digestion buffer and digested with 200 ng Trypsin (MS-Grade, Sigma Aldrich) for 19 hours at 37°C, shaking at 750 RPM. Digestion was stopped by adding 2% trifluoroacetic acid (TFA) to a final concentration of 1%. The resulting peptides were desalted on a SOLAµ SPE plate (HRP, Thermo). Between each application, the solvent was spun through by centrifugation at 350 RCF. For each sample, the filters were activated with 200 µL of 100% Methanol, then 200 µL of 80% Acetonitrile, 0.1% formic acid. The filters were equilibrated twice with 200 µL of 1% TFA, 3% Acetonitrile, after which the sample was loaded. After washing the tips twice with 200 µL of 0.1% formic acid, the peptides were eluted into clean Eppendorf Protein LoBind tubes using 40% Acetonitrile, 0.1% formic acid. The eluted peptides were concentrated in an Eppendorf Speedvac and reconstituted in 50 mM HEPES pH 8.5 for TMT labeling with 16plex tags (Thermo). Labeling was done according to the manufacturer's instructions, and subsequently pooled in two separate TMT pools according to sample type. TFA was added to acidify and bring acetonitrile concentration down to < 5%. Prior to mass spectrometry analysis, the peptides were desalted following the same procedure as described previously and fractionated using an offline ThermoFisher Ultimate3000 liquid chromatography system using high pH fractionation (5 mM Ammonium Bicarbonate, pH 10) at 5 µL/min flow rate. 15 µg of each sample pool were separated over a 120-minute gradient (5% to 35% Acetonitrile), while collecting fractions every 130 seconds. The resulting 40 fractions were pooled into 20 final fractions, acidified to pH < 2 with 1% TFA and loaded onto EvoSep stage tips according to the manufacturer's protocol.

**MS analysis** For each fraction, peptides were analysed using the pre-set '20 samples per day' method on the EvoSep One instrument. Peptides were eluted over a 58-min gradient, and analysed with an Orbitrap EclipseTM TribridTM instrument (Thermo Fisher Scientific) with FAIMS ProTM Interface (ThermoFisher Scientific) switched between CVs of -50 V and -70 V with cycle times of 1.5 s. Full MS spectra were collected at a resolution of 120,000, with normalized AGC target set to 'standard' or maximum injection time of 50 ms and a scan range of 375–1500 m/z. MS1 precursors with an intensity of >5×103 and charge state of were selected for MS2 analysis. Dynamic exclusion was set to 60 s, the exclusion list was shared between CV values and Advanced Peak Determination was set to ‘off’. The precursor fit threshold was set to 70% with a fit window of 0.7 m/z for MS2. Precursors selected for MS2 were isolated in the quadrupole with a 0.7 m/z window. Ions were collected for a maximum injection time of 50 ms and normalized AGC target set to ‘standard’. Fragmentation was performed with a CID normalised collision energy of 35% and MS2 spectra were acquired in the IT at scan rate rapid. Precursors were subsequently filtered with an isobaric tag loss exclusion of TMT and precursor mass exclusion set to 18 m/z low and 5 m/z high. Precursors were isolated for an MS3 scan using the quadrupole with a 2 m/z window, and ions were collected for a maximum injection time of 86 ms and normalized AGC target of 200%. Turbo TMT was deactivated and number of dependent scans set to 5. Isolated precursors were fragmented again with 63% normalised HCD collision energy, and MS3 spectra were acquired in the orbitrap at 50000 resolution with a scan range of 100-500 m/z. MS performance was verified for consistency by running complex cell lysate quality control standard.

**Data analysis** The raw files were analysed using Proteome Discoverer 2.4 (Thermo Fisher Scientific). TMT reporter ion quantitation was enabled in the processing and consensus steps, and spectra were matched against the Mus Musculus database obtained from UniProt. Dynamic modifications were set as Oxidation (M), and Acetyl on protein N-termini. Cysteine carbamidomethyl (C) and TMT 16-plex (peptide N-termini and K) were set as static modifications. All results were filtered to a 1% FDR, and protein quantitation done using the built-in Minora Feature Detector with statistical significance testing done with the built-in t test. The peptide abundances are normalized based on total peptide amount. Thereby, the total sum of identified peptides in a channel is normalized to the channel with the highest abundance. The protein or peptide abundances are normalized to total peptide abundance.
